# A Prospective Study to Improve Automated Blood Culture Diagnostics in Gram-Negative Sepsis by Implementing a “Diagnostic Stewardship Care-Bundle”

**DOI:** 10.1101/2022.04.22.22274034

**Authors:** Ayush Gupta, Farha Siddiqui, Shashank Purwar, Saurabh Saigal, Jai Prakash Sharma, Sanjeev Kumar

## Abstract

**Objectives:** We implemented a diagnostic stewardship care-bundle (DSB): “Sepsis-48” with the aim of evaluating its impact on changes in duration of key steps in automated blood culture diagnostics (aBCD), compliance to care-bundle and turnaround time (TAT).

**Methods:** In this prospective study, a care-bundle checklist for automated blood culture bottles (BCBs) received from adult intensive care units (AICUs) was implemented between July 2020-June 2021 (intervention period, P2) and compared with a retrospective, pre-intervention period (P1) between March-June 2020. Microbial identification in positive BCBs with gram-negatives (+nBCBs) was enabled by inoculating conventional biochemical tests directly (dID) and direct antimicrobial susceptibility testing (dAST) was done as per EUCAST RAST standard. Clinical reports were issued only if a RAST reportable gram-negative was identified in dID.

**Results:** We observed significant reductions in the Loading time (LT) [63.5 (104.5-24.5) vs 32 (55-14) minutes, P<0.001] & time to dID+dAST performance (TTD) [186 (288.25-202.25) vs 115 (180-68) minutes, P=0.0018] in +nBCBs received from AICUs during P2. There was a significant increase in compliance to the bundle targets [LT≤45: 44% vs 66%, P=.006 and TTD≤120: 34% vs 51.7%, P=.03] during P2 (Table 2). Using dID+dAST method, provisional results could be read ∼13 hours earlier than those generated by VITEK^®^. Similar improvements were also noted for +nBCBs received from other locations.

**Conclusions:** The “diagnostic stewardship care-bundle” strategy to improve aBCD was successfully implemented leading to significant reductions in duration of targeted steps. Laboratories should implement “diagnostic stewardship care-bundles” as per their needs to improve microbiological diagnostics.

## INTRODUCTION

Sepsis is one of the leading causes of mortality worldwide with highest burden in low- and middle-income countries (LMICs) [1]. It was estimated that 49 million cases occurred globally in 2017, causing 11 million deaths [2]. Sepsis mortality reflects inadequacy in healthcare infrastructure, suboptimal quality of care, poor compliance to infection prevention measures, delay in diagnosis, and inappropriate clinical management [1]. There is 9% increase in odds of mortality with each hour delay in institution of appropriate antimicrobials in sepsis [3]. Blood cultures are the cornerstone for microbiological diagnosis but with the available microbiological tools in LMICs, turnaround time (TAT) of microbiologically positive blood culture reports with antimicrobial susceptibility testing (AST) results is around 48-72 hours even with automated systems. This TAT is considered subpar with the demands of modern healthcare [4] consequently, broad-spectrum antimicrobials are used empirically with impunity creating a global pandemic of antimicrobial resistance. Hence, there is a dire need to reduce TAT of automated blood culture diagnostics (aBCD) to improve patient outcomes in sepsis and reduce broad-spectrum antimicrobial use.

Diagnostic stewardship (DS) is an important tool to improve microbiological diagnostics and can be practiced efficiently using “care-bundle approach”. It emphasizes on timely and accurate delivery of data to clinicians for efficient patient management [5]. A bundle is a set of 3-5 evidence-based interventions which when implemented together, result in significantly better outcomes than when implemented individually [6]. We conducted this study with the aim of evaluating the impact of an indigenously developed diagnostic stewardship care-bundle (DSB) to improve aBCD in our setting.

## METHODS

### Setting & Study Design

This prospective, quasi-experimental study was conducted in a continuously operational bacteriology laboratory of an academic, 950 bedded, tertiary care hospital in India between July 2020-June 2021, intervention-period (P2). The laboratory is equipped with BacT/Alert^®^ -3D (bioMerieux, Marcy d’Etoille, France) continuous blood culture monitoring system (CBCMS) for processing of automated blood culture bottles (BCBs) [FA Plus^®^ and PF Plus ^®^ (bioMerieux, Marcy d’Etoille, France)] & VITEK^®^ -2 Compact system (bioMerieux, Marcy d’Etoille, France) for microbial identification and AST. The routine operational hours are from 9 am – 5 pm and emergency hours being 5 pm – 9 am, wherein a laboratory technician is physically present to process samples and an on-call resident to report gram-stain from positively-flagged BCBs (+BCBs). The standard-of-care (SoC) method for +BCBs is subculturing on conventional plated media namely Chocolate agar, Blood agar & MacConkey agar (Himedia^®^, Mumbai, India), incubating aerobically for 16-24 hours followed by identification (GNID/GPID^®^ card) and AST (N280/281^®^ card) by VITEK^®^ once isolated colonies appear. Identification of organisms directly from +BCBs (dID) and AST testing (dAST) was implemented since January 2020 for provisional reporting (pBCR) of +BCBs with mono-morphic gram-negative bacilli or cocco-bacilli (+nBCBs). All other +BCBs underwent processing by SoC method only.

### Intervention

We developed a checklist based on “Sepsis-48” DSB (Table 1), as a quality improvement initiative with the aim of providing pBCR in BCBs, received from AICUs, within 48-hours of sample receiving. Briefly, the care-bundle targeted to optimize the key preanalytical and analytical parameters in aBCD namely; achieving loading time (LT) within 45-minutes of sample receiving, reporting gram-stain (TTG) within 60-minutes of positive signal, performing direct identification (dID) and direct antimicrobial susceptibility testing (dAST) from +nBCBs as per European Committee on antimicrobial susceptibility testing (EUCAST) rapid antimicrobial susceptibility testing (RAST) standard [7] within 120-minutes of positive signal (TTD) and reading dID+dAST results (TTR) within 48-hours of sample receiving. Personnel working in the laboratory were trained to perform, read, and report by dID+dAST method and instructed to adhere to timelines as per a checklist based on Sepsis-48 DSB, attached to requisition forms of BCBs received from AICUs only.

**Table 1:**
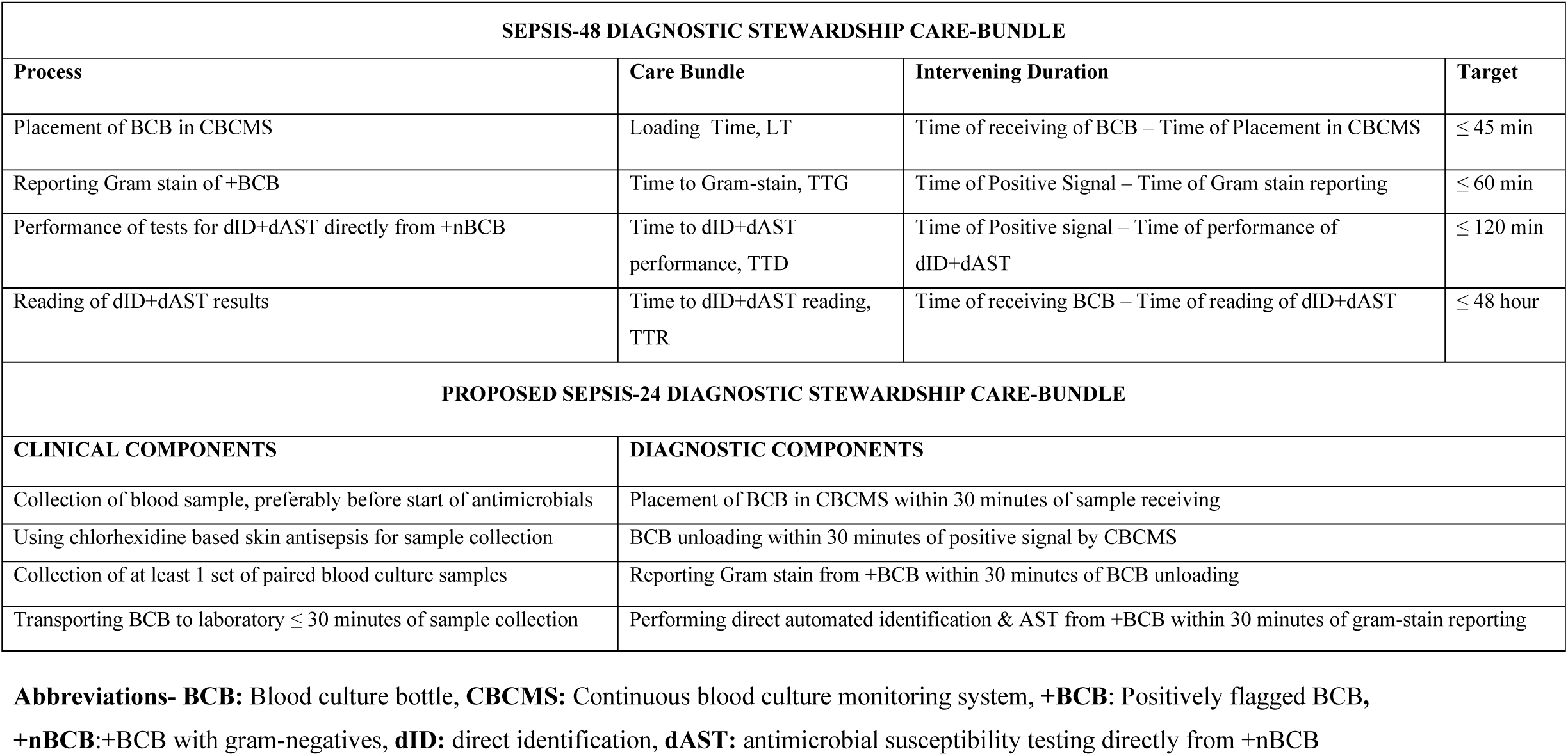
Diagnostic stewardship care-bundles

### Data collection

Time-points of various steps in aBCD were noted for +nBCBs during P2. Of them, time of sample receiving, gram-stain reporting, performance and reading of dID+dAST results and reading of final results by SoC were noted manually whereas, time of BCB loading, positive signal in CBCMS and VITEK^®^ result completion was noted from the respective automated systems. Intervening durations namely: LT, time to positivity (TTP), TTG and TTD were calculated for all +nBCBs received during study periods. TTR, time to VITEK completion (TTV) and time to final report (TTFR) were calculated for concordant +BCBs where pBCR was released. The corresponding durations in a pre-intervention period (P1) from March-June 2020, were compared to those in Intervention period, P2. Adherence to compliance with the bundle and its components was determined from the checklist.

### Microbiological Methods

For dID, microbial identification was enabled by inoculating conventional biochemical tests from the pellet obtained from positive blood-broth mixture; reading and reporting of which is described in Table 2 [8]. For dAST, EUCAST RAST method [7] was followed and results at 8-hour incubation were used for reporting [9]. Briefly, 125±25 µl of blood-broth inoculum was aspirated in 1 ml syringe and lawn culture was done on Mueller-Hinton agar (MHA, Himedia^®^, Mumbai, India) plates for dAST. Antibiotic discs (potency in µg, Himedia^®^, Mumbai, India) were applied to streaked plates and incubated subsequently at 35±2°C, namely: piperacillin-tazobactam (30-6), ceftazidime (10), meropenem (10), imipenem (10), ciprofloxacin (5), levofloxacin (5), amikacin (10), gentamicin (10), and trimethoprim-sulphamethoxazole (1.25-23.75). Provisional reports of dID+dAST method (pBCRs) were released only in mono-morphic RAST reportable gram-negatives i.e., *Escherichia coli, Klebsiella pneumoniae, Acinetobacter baumannii* complex or *Pseudomonas aeruginosa*. Isolates showing any other biochemical patterns in dID, were interpreted as “unidentified” and dAST results were not reported. Isolates which were not tested by VITEK^®^ in SoC were excluded. For dAST, weekly quality control testing was performed with *E. coli* ATCC^®^ 25922 as per RAST standard [7].

**Table 2:** Performance and interpretation of direct microbial identification (dID) method by directly inoculating conventional biochemical tests

| Biochemical Test | Time of Reading, Post-inoculation (hrs) | *Escherichia coli* | *Klebsiella pneumoniae* | *Pseudomonas aeruginosa* | *Acinetobacter baumannii* complex |
| --- | --- | --- | --- | --- | --- |
| Colonies on MA | 18-24 | LF/NLF | LF | NLF | NLF/LLF |
| Catalase | 8 | Positive | Positive | Positive | Positive |
| Oxidase | 8 | Negative | Negative | Positive | Negative |
| Motility by Hanging drop | 8 | M/NM | NM | M | NM |
| Indole test | 18-24 | Positive | Negative | Negative | Negative |
| Simmon's Citrate agar | 18-24 | Negative | Positive | Positive | Positive |
| Christensen's Urease agar | 18-24 | Negative | Positive/<br>Negative | Negative | Negative |
| Mannitol Motility agar | 18-24 | F,<br>M/NM | F,<br>NM | NF,<br>M | NF,<br>NM |
| Triple Sugar Iron agar | 18-24 | A/A or K/A,<br>no H<sub>2</sub>S | A/A, no H<sub>2</sub>S | K/No change | K/No change |
| Hugh-Leifson's OF test | 18-24 | Fermentative | Fermentative | Oxidative | Oxidative |

**Abbreviations:** MA: MacConkey agar, LF: Lactose Fermenter, NLF: Non-Lactose Fermenter, LLF: Late Lactose Fermenter, M: Motile, NM: Non motile, F: Fermenter, NF: Non fermenter, A: Acid, K: Alkaline, H<sub>2</sub>S: Hydrogen sulphide gas, OF: Oxidation-Fermentation

### Statistical analysis

Result of microbial identification by VITEK^®^ was considered as “gold standard” and +nBCBs with same interpretation (till species-level of RAST reportable gram-negatives) in dID, were considered as “concordant”. Positive predictive value (PPV) of dID method was calculated as proportion of concordant results to total number of dID+dAST reports released. Negative predictive value (NPV) was calculated as proportion of results correctly interpreted as “unidentified” by dID to total number of results interpreted as “unidentified” in dID.

For AST, results by VITEK^®^ N280/281^®^ cards were considered as “gold standard”. Results categorized under “area of technical uncertainty (ATU)” in dAST or VITEK^®^ were excluded from analysis as standards recommends against their reporting [9]. Results were in categorical agreement (CA) when results of a particular drug-bug combination were same by both dAST and VITEK^®^. Disagreements were further categorized as major error rate (ME%= resistant, R by dAST*100/S, susceptible by VITEK^®^); very major error rate (VME%), if opposite of ME% and as minor error (mE) when showing any other pattern [10]. Percentages (%) for CA & mE were calculated by multiplying the ratio of total number in respective category (numerator) by total number of such drug-bug combinations tested *sans* ATU (denominator) by 100 [10]. PPV (S by dAST*100/ S by both VITEK^®^ and dAST) & NPV of dAST (non-susceptible, NS by dAST*100/ NS by both VITEK^®^ and dAST) were also calculated [11].

Data was managed and analysed using Microsoft^®^ Excel 365. Mann-Whitney U and Chi-square (χ^2^) tests were used for continuous and categorical variables, respectively considering a two-sided p-value of ≤0.05 as statistically significant.

## RESULTS

### Subjects

During P1 & P2, 1,028 and 5,016 BCBs respectively, were received from all locations. Of these, 26.9% (277/1028) and 28.1% (1409/5016) were from AICUs. Details of cohort assembly in each period is provided in Fig.1. The differences in the general characteristics of BCBs received during the two periods is shown in Table 3. Briefly, BCBs during P2 had less proportion of +nBCB and significantly higher number of RAST reportable gram-negatives. Overall, dID+dAST was performed in 11.7% (120/1028) and 7.7% (385/5016) +nBCBs during P1 & P2, respectively of which proportion of +nBCBs from AICUs was 41.7% (50/120) and 38.2% (147/385), respectively. Overall, 32 concordant dID+dAST reports were released in +nBCBs received from all locations during P1 of which 12 were from AICUs whereas the corresponding figures during P2 were 133 and 57 during P2.

**Figure 1:**
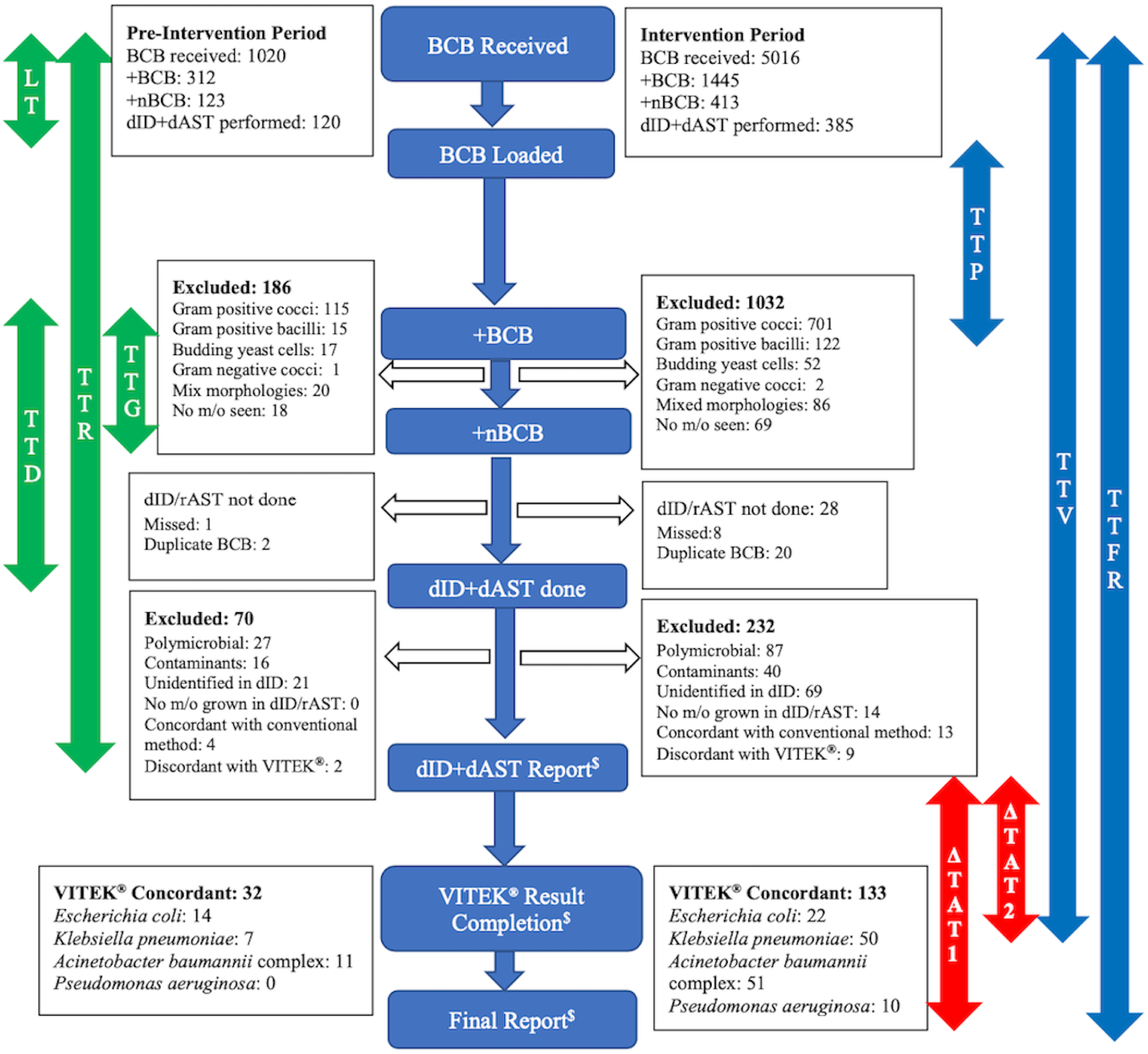
Summary of cohort assembly in the study^#^ **Abbreviations**: **BCB:** automated blood culture bottles, +BCB: Positively flagged BCB, +nBCB: +BCB with gram-negatives, **m/o:** micro-organisms, **dID:** direct identification, **dAST:** direct antimicrobial susceptibility testing from +nBCB, **LT:** loading Time, **TTG:** time to Gram stain, **TTD:** time to dID+dAST performance, **TTR:** time to dID+ dAST reading, **TTP:** time to positivity, **TTV:** time to VITEK results, **TTFR:** time to final report by standard of care method, **ΔTAT1**: time difference between TTFR & TTR, **ΔTAT2**: time difference between TTV & TTR # LT, TTG & TTD were calculated for 385 BCBs for which dID+dAST was done, whereas TTR, TTV & TTFR were calculated for 133 VITEK^**®**^ concordant +nBCBs, for which provisional blood culture reports (pBCRs) were released

**Table 3:** Comparison of general characteristics of automated blood culture bottles received during both periods

| GENERAL CHARACTERISTICS | PRE-INTERVENTION | INTERVENTION | P-VALUE<sup>1</sup> |
| --- | --- | --- | --- |
| BCB Received | 1028 | 5016 |  |
| AICUs | 26.9% (277/1028) | 28.9% (1449/5016) | .20 |
| Other Locations | 73.1% (751/1028) | 71.1% (3567/5016) |  |
| +BCB | 30.3% (312/1028) | 28.8% (1445/5016) | .32 |
| +nBCB | 39.4% (123/312) | 28.6% (413/1445) | **.0001<sup>2</sup>** |
| +pBCB | 36.8% (115/312) | 48.5% (701/1445) | **.0001** |
| +oBCB | 23.7% (74/312) | 23.6% (341/1445) | .96 |
| dID+dAST done | 97.6% (120/123) | 93.2% (385/413) | .07 |
| AICUs | 41.7% (50/120) | 38.2% (147/385) | .49 |
| Other Locations | 58.3% (70/120) | 61.8% (238/385) |  |
| Time of BCB receiving |  |  |  |
| Routine hours | 67.5% (81/120) | 61% (235/385) | .20 |
| Emergency hours | 32.5% (39/120) | 39% (150/385) |  |
| Time of BCB Signalling |  |  |  |
| Routine hours | 26.7% (32/120) | 30.9% (119/385) | .37 |
| Emergency hours | 73.3% (88/120) | 69.1% (266/385) |  |
| Concordant dID+dAST reported | 26.7% (32/120) | 34.5% (133/385) | .10 |
| AICUs | 37.5% (12/32) | 42.8% (57/133) | .58 |
| Other Locations | 62.5% (20/32) | 57.2% (76/133) |  |
| RAST Reportable Micro-organisms in all +nBCBs | 28.3% (34/120) | 38.2% (147/385) | **.04** |
| *Escherichia coli* | 44.1% (15/34) | 17.7% (26/147) | **.0009** |
| *Klebsiella pneumoniae* | 20.6% (7/34) | 36.7% (54/147) | .07 |
| *Acinetobacter baumannii* complex | 35.3% (12/34) | 37.4% (55/147) | .81 |
| *Pseudomonas aeruginosa* | 0 | 8.2% (12/147) | .12 |

**Abbreviations:** BCB: automated blood culture bottle, AICUs: adult intensive care units, +BCB: positively flagged BCB, +nBCB: +BCB with gram-negatives, +pBCB: +BCB with gram positive cocci, +oBCB: +BCB with other results in gram-stain (mix morphology, gram positive bacilli, budding yeast cells and no micro-organisms seen, dID: direct microbial identification, dAST: direct antimicrobial susceptibility testing, RAST: Rapid antimicrobial Susceptibility testing method by EUCAST
<sup>1</sup> Chi-square or Fischer's exact test; <sup>2</sup> Significant p-values are in bold

### Microbiological results

A total of 505 dID+dAST were performed during both study periods of which 120 were done during P1 and 385 during P2. Amongst them, 41.7% (50/120) and 38.2% (147/385), respectively were from AICUs. Overall, 176 dID+dAST reports, having a RAST reportable gram-negative i.e., either *E. coli, K. pneumoniae, A. baumannii* complex or *P. aeruginosa* were released during both the study periods of which 93.7% (165/176) were concordant with VITEK^®^. By using dID method, we failed to identify one of the RAST reportable gram-negative in 14 +nBCBs amongst 90 monomorphic +nBCBs which were reported as “unidentified. Details of the reporting errors have been described in Table 4. The PPV of dID, till species- and genus-level identification, were 93.7% (165/176) and 96% (169/176), respectively and NPV was 84.4% (76/90).

**Table 4:** Results of direct microbial identification method by utilizing conventional biochemical tests

| Wrong dID+dAST Reports Sent |  |  | No dID+dAST Reports Sent |  |  |
| --- | --- | --- | --- | --- | --- |
| Result by dID | Result by VITEK® | Number | Result by dID | Result by VITEK® | Number |
| *Acinetobacter baumannii* complex | *Achromobacter xylosoxidans* | 1 | Unidentified | *Escherichia coli* | 4 |
| *Acinetobacter baumannii* complex | *Acinetobacter lwoffii* | 2 | Unidentified | *Klebsiella pneumoniae* | 4 |
| *Acinetobacter baumannii* complex | *Escherichia coli* | 1 | Unidentified | *Acinetobacter baumannii* complex | 5 |
| *Acinetobacter baumannii* complex | *Pseudomonas aeruginosa* | 1 | Unidentified | *Pseudomonas aeruginosa* | 1 |
| *Acinetobacter baumannii* complex | *Burkholderia cepacia* complex | 1 | Unidentified | *Achromobacter denitrificans*: 2<br>*Achromobacter xylosoxidans*: 2<br>*Acinetobacter lwoffii*: 5<br>*Acinetobacter radioresistans*: 7<br>*Aeromonas salmonicida*: 1<br>*Brevundimonas diminuta*: 1<br>*Burkholderia cepacia* complex: 4<br>*Burkholderia pseudomallei*: 4<br>*Chryseobacterium indologenes*: 2<br>*Elizabethkingia meningoseptica*: 2<br>*Enterobacter aerogenes*: 1<br>*Enterobacter cloacae* complex: 6<br>*Moraxella* spp.: 1<br>*Ochrobactrum anthropi*: 1<br>*Pantoea agglomerans*: 1<br>*Pseudomonas putida*: 1<br>*Raoultella ornitholytica*: 1<br>*Salmonella* Paratyphi A: 1<br>*Salmonella* Typhi: 12<br>*Serratia marcescens*: 2<br>*Serratia odorifera*: 1<br>*Sphingomonas paucimobilis*: 5<br>*Stenotrophomonas maltophilia*: 13 |  |
| *Klebsiella pneumoniae* | *Enterobacter cloacae* complex | 2 |  |  |  |
| *Klebsiella pneumoniae* | *Sphingomonas paucimobilis* | 1 |  |  |  |
| *Klebsiella pneumoniae* | *Klebsiella oxytoca* | 1 |  |  |  |
| *Pseudomonas aeruginosa* | *Pseudomonas putida* | 1 |  |  |  |
| Concordant Identifications |  |  |  |  |  |
| *Escherichia coli* |  | 36 |  |  |  |
| *Klebsiella pneumoniae* |  | 57 |  |  |  |
| *Acinetobacter baumannii* complex |  | 62 |  |  |  |
| *Pseudomonas aeruginosa* |  | 10 |  |  |  |

**Abbreviations:** dID: direct microbial identification, dAST: direct antimicrobial susceptibility testing

Amongst 165 concordant +nBCBs during both study periods, 62 (37.6%) were *A. baumannii* complex, 57 were *K. pneumoniae* (34.5%), 36 (21.8%) were *E. coli* and 10 (6.1%) were *P. aeruginosa*. Their antimicrobial susceptibility patterns are provided in Fig. 2. Overall, 1,237 drug-bug combination were tested in dAST, of which 73 (5.9%) were in “ATU” category and were excluded from further analysis. Overall, CA for any drug-bug combination was 92.7% (1079/1164) and corresponding ME, VME and mE rates were 8.8% (19/216), 4.9% (45/921) and 1.8% (21/1164), respectively. The corresponding rates for each tested drug-bug combination, PPV and NPV are given in Table 5.

**Figure 2:**
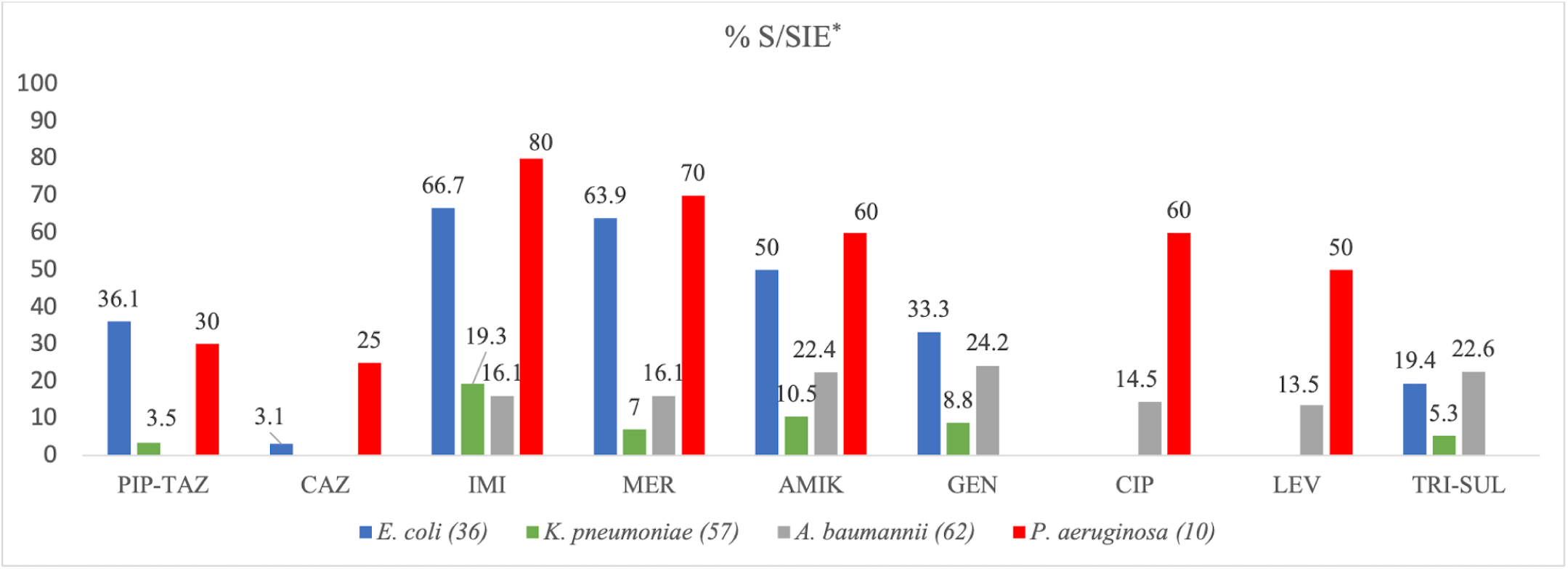
Antimicrobial Susceptibility pattern of Isolates included in the analysis (As per VITEK^®^-2 Compact system, interpreted as per EUCAST breakpoint tables for full incubation, version: 10) **Abbreviations:** S: susceptible, SIE: susceptible, increased exposure, PIP-TAZ: piperacillin-tazobactam, CAZ: ceftazidime, IMI: imipenem, MER: meropenem,, AMIK: amikacin, GEN: gentamicin, CIP: ciprofloxacin, TRI-SUL: trimethoprim-sulphamethoxazole, *E: Escherichia, K: Klebsiella, A: Acinetobacter, P: Pseudomonas* **\*** Susceptible, increased exposure interpretive category is applicable for piperacillin-tazobactam, ceftazidime, imipenem, ciprofloxacin and levofloxacin against *P. aeruginosa* and ciprofloxacin against *A. baumannii* complex **Note:** No interpretive criteria available for piperacillin-tazobactam and ceftazidime against *A. baumannii* complex and for gentamicin and trimethoprim-sulphamethoxazole against *P. aeruginosa*

**Table 5:**
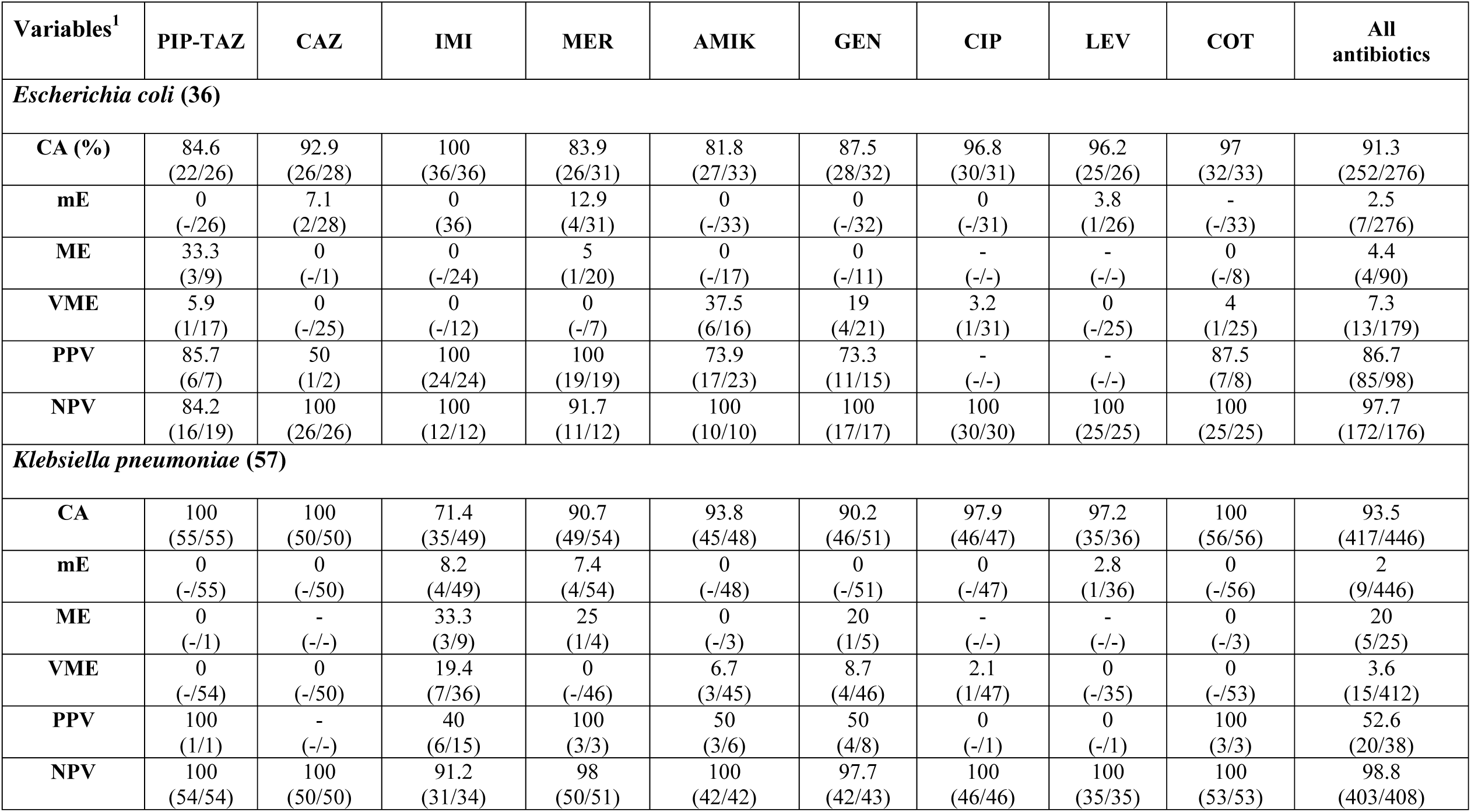
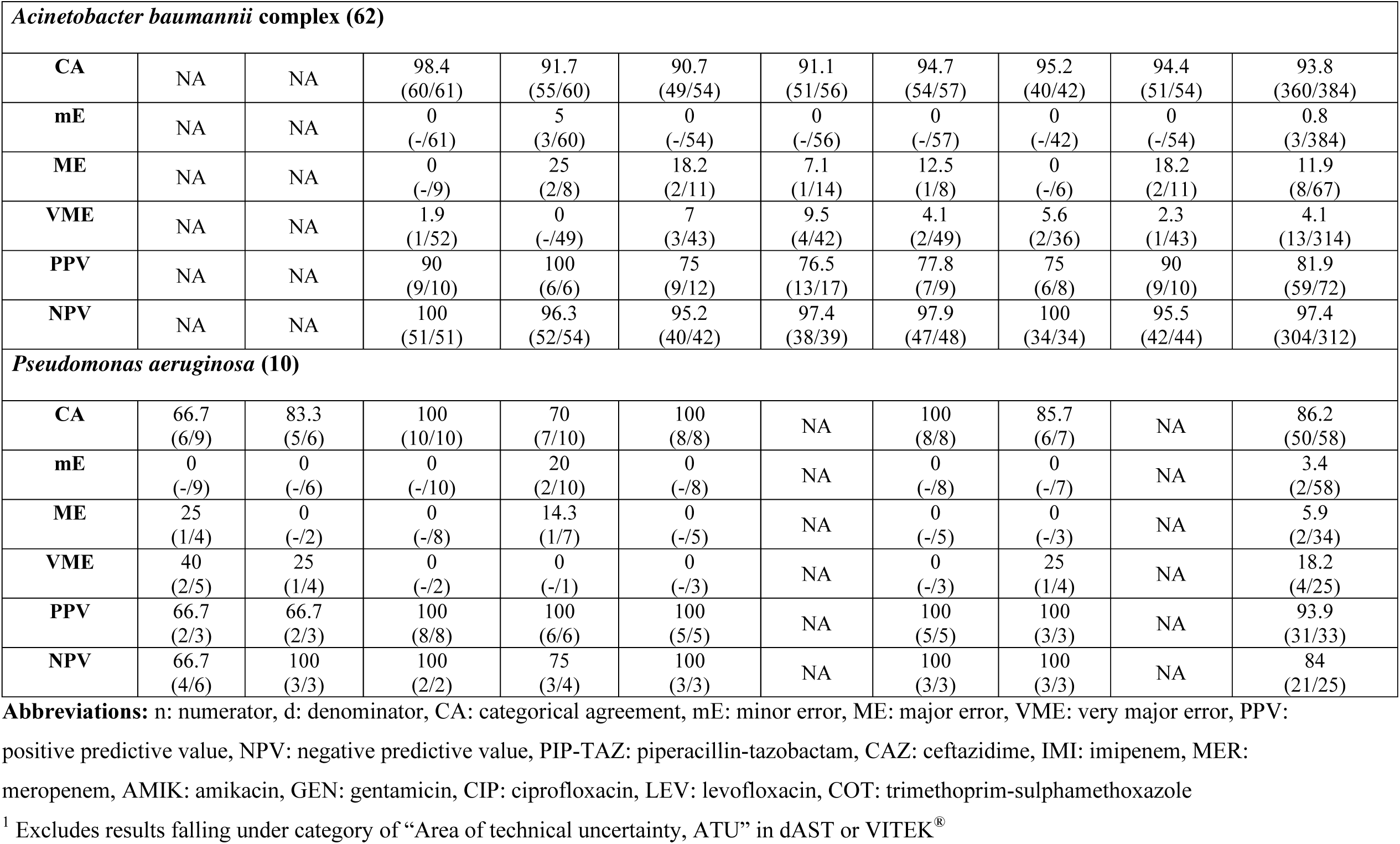
Diagnostic accuracy (CA, mE, ME and VME) and predictive values (PPV and NPV) of rAST method in comparison to antimicrobial susceptibility testing by VITEK^®^ (interpreted as per EUCAST breakpoints for full incubation)

### Impact of Sepsis-48 DSB

LT, TTP, TTG and TTD were calculated for 120 and 385 +nBCBs received during P1 and P2, respectively whereas TTR, TTV and TTFR were calculated for 32 and 133 concordant identifications, respectively. The corresponding changes in duration of these pre- and analytical parameters amongst +nBCBs received from all locations, and subset analysis between AICUs and other locations is shown in Table 6. Similarly, the changes in adherence to compliance with the components of care bundles are shown in Table 6. Notably, “all-or-none” compliance to the first 3 components of DSB i.e., LT≤45, TTG≤60 and TTR≤120 increased significantly from 10.8% (13/120) in P1 to 22.1% (85/385) BCBs (p=.006, χ^2^-test) in P2.

**Table 6:** Impact of “Sepsis-48” diagnostic stewardship bundle on various time intervals and compliance to specified care bundles.

|  | DURATION |  |  | COMPLIANCE |  |  |
| --- | --- | --- | --- | --- | --- | --- |
|  | PRE-INTERVENTION<br>Median (IQR), minutes | INTERVENTION<br>Median (IQR), minutes | P-VALUE<sup>1</sup> | PRE-INTERVENTION<br>Compliance % (n/d) | INTERVENTION<br>Compliance % (n/d) | P-VALUE<sup>2</sup> |
| **ALL LOCATIONS** |  |  |  |  |  |  |
| **LT ≤45 min** | 64.5 (103.5 – 23.5) | 32 (59–14) | **< .001** | 44.2 (53/120) | 64.1 (247/385) | **< .001<sup>3</sup>** |
| **TTP** | 990 (1298.75-701) | 1001 (1360.75-772.5) | .27 | - | - |  |
| **TTG ≤60 min** | 68 (149.75 – 39.25) | 63 (119 – 30) | .15 | 45 (54/120) | 49.3 (190/385) | 0.40 |
| **TTD ≤120 min** | 186 (302.75 - 92) | 128 (207.5 - 78.5) | **< .001** | 33.3 (40/120) | 47.8 (184/385) | **0.005** |
| **TTR ≤48 hr** | 2535 (2791 - 2263.5) | 2540 (2744 - 2218) | .77 | 84.4 (27/32) | 85.7 (114/133) | .84 |
| **TTV** | 3386.5 (3520 - 3119) | 3234 (3493.5 - 2907) | .06 | - | - |  |
| **TTFR** | 4086 (4234.5-3750) | 3970 (4222.5-3620) | .34 | - | - |  |
| **Δ TAT2** | 694 (1054.5-617.25) | 646 (806.5-441.5) | .10 | - | - |  |
| **Δ TAT1** | 1400 (1500-1440) | 1440 (1500-1140) | .25 |  |  |  |
| **ADULT ICUs** |  |  |  |  |  |  |
| **LT ≤45 min** | 63.5 (104.5 – 24.5) | 32 (55 – 14) | **< .001** | 44 (22/50) | 66 (97/147) | **.006** |
| **TTP** | 907.5 (1185.75-676.75) | 970 (1274-773) | .19 | - | - |  |
| **TTG ≤60 min** | 72 (180 – 31.75) | 51 (104 – 30) | .06 | 44 (22/50) | 56.5 (83/147) | .12 |
| **TTD ≤120 min** | 186 (288.25 – 202.25) | 115 (180 - 68) | **.0018** | 34 (17/50) | 51.7 (76/147) | **.03** |
| **TTR ≤48 hr** | 2500 (2787.5 – 2347.5) | 2550 (2765 – 2260) | .88 | 83.3 (10/12) | 84.2 (48/57) | .93 |
| **TTV** | 3308.5 (3662.2 – 3097.2) | 3268 (3504.5 – 2909.5) | .51 | - | - |  |
| **TTFR** | 3940 (4238.75-3817.5) | 4020 (4250-3695) | .83 | - | - |  |
| **Δ TAT2** | 694 (841.75-625.75) | 652 (782.5-396) | .39 | - | - |  |
| **Δ TAT1** | 1440 (1451.25-1440) | 1440 (1485-1110) | .53 | - | - |  |

| OTHER LOCATIONS |  |  |  |  |  |  |
| --- | --- | --- | --- | --- | --- | --- |
| **LT ≤45 min** | 66.5 (106.2 – 20.5) | 32 (61.25 – 14) | **.001** | 44.3 (31/70) | 62.6 (149/238) | **.006** |
| **TTP** | 1013.5 (1346.25-728.25) | 1022.5 (1468.5-770.25) | .72 | - | - |  |
| **TTG ≤60 min** | 66 (130-.5 – 40.7) | 70 (127.2 – 33) | .81 | 45.7 (32/70) | 45.4 (108/238) | .96 |
| **TTD ≤120 min** | 186 (312 – 96) | 134.5 (215.2 – 87.7) | **.009** | 32.3 (23/70) | 45 (107/238) | .07 |
| **TTR ≤48 hr** | 2577.5 (2791 – 2100) | 2512.5 (2727.5 – 2159.2) | .77 | 85 (17/20) | 86.8 (66/76) | .73 |
| **TTV** | 3386.5 (3520 – 3325) | 3213.5 (3490.25 -2833.5) | .07 | - | - |  |
| **TTFR** | 4122.5 (4234.5-3660) | 3960 (4200-3570) | .28 | - | - |  |
| **Δ TAT2** | 709 (1209.75-562.75) | 616 (848-446.75) | .21 | - | - |  |
| **Δ TAT1** | 1440 (1552.5-1256.25) | 1440 (1500-1140) | .35 | - | - |  |

**Abbreviations:** **IQR:** Interquartile range, **n:** numerator, **d:** denominator, **LT:** Loading time, **min:** minutes, **TTG:** Time to gram-stain, **TTD:** Time to dID+dAST performance, **TTR:** Time to dID+dAST reading, **hr:** hour, **TTV:** Time to VITEK<sup>®</sup> result completion, **TTFR:** Time to release of final report by standard of care method, **Δ TAT2:** TAT difference between VITEK<sup>®</sup> result completion (TTV) and dID+dAST report (TTD), **Δ TAT1:** TAT difference between Final report (TTFR) and dID+dAST report (TTD), **ICUs:** Intensive care units
<sup>1</sup> Mann-Whitney U-test, <sup>2</sup> Chi-square test, <sup>3</sup> Significant p-values are in bold

## DISCUSSION

Care-bundle approach has shown to improve practices in infection control [12] and antimicrobial stewardship [13] however, evidence on the impact of various diagnostic stewardship interventions is limited [14]. Our intent while designing the bundle was to affect processes which can minimize delays in clinical reporting of aBCD. While designing it, we faced difficulty to determine the evidence on recommended durations of various key steps in aBCD. For example, one can find literature on the impact of immediate gram-staining, but it is difficult to measure “what impact does reporting gram-stain from a +BCB within 60 minutes of positive signal have on patient outcome?”. Hence, the components of the care-bundle were selected on the merit of “what constitutes as best practice” and key reasons for diagnostic delays in our setting.

We found that by implementing the care-bundle during P2, there were significant reductions in durations of LT and TTD and increase in compliance to LT ≤45 and TTD ≤120 minutes in +nBCBs from AICUs (Table 6). Similar improvements were also noted in +nBCBs from other locations which underscores the importance of percolation of quality improvement initiatives in overall practices of the system (Table 6). The care-bundle couldn’t significantly reduce TTG however, it reduced by 29% from 72 to 51 minutes in +nBCBs from AICUs only (p=.06). This was because ∼54% of +nBCBs gave signal between 8 pm-8 am and resident doctors had to travel before reporting. On subset analysis, we found that the TTG during emergency hours improved (100 vs 65 minutes, P1 vs P2, p=.06) whereas it reduced marginally from 36 to 32 minutes during routine hours.

We found that the care-bundle couldn’t reduce the overall duration of reporting of either dID+dAST or SoC method, during P2 (Table 6). This was multifactorial, firstly, during aBCD, ∼80% component of TAT is a function of bacterial growth including TTP and duration of incubation for identification and susceptibility tests, which were not targeted in the bundle. The major reason for reduction of TAT in pBCR was due to direct identification and susceptibility tests which were already being performed during P1. Hence, it was unreasonable to judge the impact of care-bundle on overall reporting times. Thirdly, the compliance rates to individual components of bundle increased moderately but the “all-or-none” compliance to bundle was poor. It is recommended that for a bundle to be effective the adherence to individual components should be ≥95% [6]. Fourthly, TTP, which is a function of rate of bacterial growth was higher during P2, offsetting any improvement in the time gained in other steps.

We performed dID+dAST method with considerable diagnostic accuracy. We correctly reported a RAST reportable gram-negative in ∼94% (165/176) and ∼96% (169/176) +nBCBs till species and genus-level, respectively. The NPV of dID method was estimated as 84% as we failed to identify a RAST reportable gram-negative in 14 of 90 +nBCBs which we considered to have a non-RAST reportable gram-negative. We found acceptable CA & mE rates in dAST method for most tested drug-bug combinations, when Cumitech defined cutoffs (CA ≥90%, VME ≤1.5% & ME ≤3%) were applied for equivalence in susceptibility testing methods [15], but VME rates were high in few combinations such as aminoglycosides against *E. coli, K. pneumoniae, A. baumannii* complex; Meropenem against *E. coli;* and Imipenem against *K. pneumoniae* (Table 5). Due to high proportion of multi-drug resistant organisms (MDROs) being isolated (∼80%) in our study, sensitive drug:bug combinations were extremely low, hence our ME rates and PPV rates are not reliable due to low denominators. Importantly, the NPV of all drug-bug combinations having ≥30 denominator was >90% implying that while a “resistant” result by dAST is highly reliable, the same cannot be said of a “sensitive” result, particularly with aminoglycosides.

There is an increasing stress on reducing TAT of aBCD, hence both Clinical Laboratory Standards Institute (CLSI) and EUCAST have developed interpretive criteria for AST directly from +BCBs [16,17]. Such rapid phenotypic method ticks all boxes for ASSURED criteria i.e., *affordable, sensitive, specific, user-friendly, rapid, equipment-free and deliverable* by WHO for a diagnostic test [18]. Due to high prevalence of MDROs in LMICs, the potential of direct AST methods will be realized in escalation of antimicrobial treatment and improvement in patient outcomes rather than as a tool for reducing antimicrobial use. The major limitation of implementing direct AST in LMICs is due to unavailability of bacterial identification results at the time of reading, which can be as early as 4-hour using EUCAST RAST method. Those implementing it in their setting have utilized matrix-assisted laser desorption/ionization time of flight mass spectrometry (MALDI-TOF MS) for microbial identification after short incubation [19–21] except in a recently published Turkish study, wherein gram-negatives were broadly categorized using simple biochemical tests like catalase, oxidase and motility. However, authors didn’t disclose the error rates with the approach [22].

We opine that it is possible to reduce TAT of aBCD further, to ≤24 hours of sample collection and propose the template of a “Sepsis-24” DSB (Table 1). It is based on the observations that mean TTP of +nBCBs is between 8-16 hours and identification of gram-negatives takes ∼4-8 hours in an automated microbial identification system. By direct inoculation of identification cards of various automated systems, studies have shown excellent concordance in identification of gram-negatives [8,23]. By utilizing the widely prevalent automated tools in LMICs and “care-bundle” approach in conjunction, this proposed DSB can act as a primer for improving aBCD in resource limited settings with minimal additional cost.

Our study also underlines the benefits of a continuously running bacteriology laboratory. We found that, in our setting, 38% (189/505) of +nBCBs were received outside of routine hours i.e., between 5 pm to 9 am and 70% (354/505) gave signal during this time duration. Due to the continuously functional laboratory, we were able to report gram-staining in emergency hours and proceed with further tests to perform direct identification and susceptibility testing in the emergency hours. As a result, we considerably reduced the TAT associated with blood culture reporting in our setting. There has been no available data in published literature on the practices related to automated blood culture diagnostics in the LMICs but even in European countries, only 13% clinical microbiology laboratories process +BCBs and 5% disseminate validated AST results to clinicians for patient management, round the clock [4]. In our opinion, round the clock functioning clinical microbiology laboratory is an important intervention to facilitate diagnostic and antimicrobial stewardship, which will have far reaching consequences in combating with the burgeoning problem of antimicrobial resistance.

Our study has several merits, firstly, the application of a targeted “care-bundle” for diagnostic stewardship in aBCD is a first and underlines the importance of employing simple practices to improve microbiological diagnostics in resource limited settings where existing infrastructure should be utilized maximally. The study was conducted prospectively in a continuously functional laboratory, with necessary tools and practices essential for implementation already in place. As a result, the difference in TATs of pBCR and reports by SoC method was around 24-hours. Thirdly, we addressed the major limitation of microbial identification with considerable diagnostic accuracy and provide a template for implementation of direct AST in settings without MALDI-TOF MS.

There were few limitations, firstly we performed the analysis only on +nBCBs, instead of all BCBs as dID+dAST was not implemented for gram-positives due to majority being non-RAST reportable. Secondly, time data of various steps in aBCD was maintained manually due to lack of a lab information system and is subject to inaccuracies while documenting. Thirdly, the “all-or-none” compliance rate to the care-bundle was poor due to poor compliance with TTG ≤60, which could have been addressed by training laboratory technicians to report gram-stain from a +BCB. Fourthly, the sample collection practices were poor leading to high rates of skin commensals and polymicrobial BCBs. As a result, we could give correct report only in one-third of all dID+dAST performed. Finally, our study was conducted in a single center only and results should be analysed in context of the prevailing micro-organisms and their antimicrobial susceptibility profile.

To conclude, there is an urgent need to provide timely results in sepsis to improve patient outcomes and reduce broad-spectrum antimicrobial use particularly in LMICs. Development of direct AST by EUCAST & CLSI is a much-needed step and ideally suited for resource limited settings if they can adapt it in their workflow. The benefits can be further augmented by using them in conjunction with “care-bundle” strategy which has been time-tested and should be explored further to practice diagnostic stewardship.

## Data Availability

All data produced in the present work are contained in the manuscript

## ETHICAL APPROVAL

For performing direct identification and susceptibility testing (dID+dAST), ethical approval was taken from Institute’s ethics committee (IEC), AIIMS Bhopal as per letter no. IHECPGRMD018 dated 11/3/2020. The implementation of diagnostic stewardship care-bundle was done as a quality improvement initiative.

## TRANSPARENCY DECLARATION

### Conflict of Interest

All authors report no conflict of interest

### Funding

The study was partially supported by the MD Thesis grant awarded to Dr. Farha Siddiqui by Indian Council of Medical Research as per letter no.:3/2/June-2020/PG-Thesis-HRD(26) dated 2/9/2020. The work has been submitted in ECCMID-2022 for oral paper/e-poster presentation.

## Acknowledgements

We wholeheartedly thank the laboratory technicians and the resident doctors who took the readings of tests, diligently at the specified time during the emergency hours.

## Access to data

Full access to the data will be available on request

## Author Contributions

Gupta A, Purwar S, Sharma JP and Saigal S conceptualized the study plan; Siddiqui F & Yadav J performed testing, acquired data and entry in software; Gupta A and Kumar S did data analysis & its interpretation; Gupta A, Siddiqui F, Yadav J and Kumar S wrote initial manuscript; Sharma JP, Saigal S & Purwar S provided critical inputs in the final manuscript. All authors approve the final draft of submitted manuscript.

## Notes

### Competing Interest Statement

The authors have declared no competing interest.

### Funding Statement

The study was partially funded by the Indian Council of Medical Research (ICMR) as a part of MD/MS Thesis Award granted to Dr. Farha Siddiqui As per letter no. 3/2/June-2020/ PG-Thesis-HRD(26) dated 2/9/20.

### Author Declarations

Institute Human Ethics Committee of All India Institute of Medical Sciences, Bhopal gave ethical approval for this work which includes performance of dID+dAST as per letter no. IHECPGRMD018 dated 11/3/2020. The implementation of diagnostic stewardship care -bundle was done as a quality improvement initiative.

## REFERENCES

[1] WHO.Global report on the epidemiology and burden of sepsis: current evidence, identifying gaps and future directions. 2020.

[2] Rudd KE, Johnson SC, Agesa KM, Shackelford KA, Tsoi D, Kievlan DR, et al. Global, regional, and national sepsis incidence and mortality, 1990–2017 : analysis for the Global Burden of Disease Study. The Lancet 2020;395:200–11. https://doi.org/10.1016/S0140-6736(19)32989-7.

[3] Liu VX, Fielding-Singh V, Greene JD, Baker JM, Iwashyna TJ, Bhattacharya J, et al. The Timing of Early Antibiotics and Hospital Mortality in Sepsis. Am J Respir Crit Care Med 2017;196:856–63. https://doi.org/10.1164/rccm.201609-1848OC.

[4] Lamy B, Sundqvist M, Idelevich EA. Bloodstream infections e Standard and progress in pathogen diagnostics. Clinical Microbiology and Infection 2020;26:142–50. https://doi.org/10.1016/j.cmi.2019.11.017.

[5] Gronthoud FA. Diagnostic Stewardship. Practical Clinical Microbiology and Infectious Diseases 2020:6–11. https://doi.org/10.1201/9781315194080-1-2.

[6] Resar R, FA G, C H, TW N. Using Care Bundles to Improve Health Care Quality. IHI Innovation Series White Paper 2012:1–14.

[7] Rast E. Methodology - EUCAST rapid antimicrobial susceptibility testing (RAST) directly from positive blood culture bottles. 2019;1.

[8] Höring S, Massarani AS, Löffler B, Rödel J. Rapid antibiotic susceptibility testing in blood culture diagnostics performed by direct inoculation using the VITEK ® -2 and BD Phoenix <sup>TM</sup> platforms 2019;2.

[9] Committee E, Testing AS. Zone diameter breakpoints for rapid antimicrobial susceptibility testing (RAST) directly from blood culture bottles European Committee on Antimicrobial Susceptibility Testing Zone diameter breakpoints for rapid antimicrobial susceptibility testing (RAS 2020:0–15.

[10] Clinical and Laboratory Standards Institute C. M52-Ed1 | Verification of Commercial Microbial Identification and Antimicrobial Susceptibility Testing Systems, 1st Edition. Clinical and Laboratory Standards Institute 2017.

[11] Savage TJ, Rao S, Joerger J, Ozonoff A, McAdam AJ, Sandora TJ. Predictive value of direct disk diffusion testing from positive blood cultures in a children’s hospital and its utility in antimicrobial stewardship. Journal of Clinical Microbiology 2021;59. https://doi.org/10.1128/JCM.02445-20.

[12] Molina García A, Cross JH, Fitchett EJA, Kawaza K, Okomo U, Spotswood NE, et al. Infection prevention and care bundles addressing health care-associated infections in neonatal care in low-middle income countries: a scoping review. EClinicalMedicine 2022;44:101259. https://doi.org/10.1016/j.eclinm.2021.101259.

[13] Panditrao A, Shafiq N, Kumar-M P, Sekhon AK, Biswal M, Singh G, et al. Impact of an antimicrobial stewardship and monitoring of infection control bundle in a surgical intensive care unit of a tertiary-care hospital in India. J Glob Antimicrob Resist 2021;24:260–5. https://doi.org/10.1016/j.jgar.2021.01.003.

[14] Bookstaver PB, Nimmich EB, Smith TJ 3rd, Justo JA, Kohn J, Hammer KL, et al. Cumulative Effect of an Antimicrobial Stewardship and Rapid Diagnostic Testing Bundle on Early Streamlining of Antimicrobial Therapy in Gram-Negative Bloodstream Infections. Antimicrob Agents Chemother 2017;61. https://doi.org/10.1128/AAC.00189-17.

[15] Clark RB, Lewinski MA, Loeffelholz MJ, Tibbetts RJ, Sharp SE. Richard b. clark, Michael a. lewinski, michael j. loeffelholz, and robert j. tibbetts. n.d.

[16] Chandrasekaran S, Abbott A, Campeau S, Zimmer BL, Weinstein M, Thrupp L, et al. Direct-from-Blood-Culture Disk Diffusion To Determine Antimicrobial Susceptibility of Gram-Negative Bacteria: Preliminary Report from the Clinical and Laboratory Standards Institute Methods Development and Standardization Working Group. J Clin Microbiol 2018;56. https://doi.org/10.1128/JCM.01678-17.

[17] Åkerlund A, Jonasson E, Matuschek E, Serrander L, Sundqvist M, Kahlmeter G. EUCAST rapid antimicrobial susceptibility testing (RAST) in blood cultures: validation in 55 European laboratories. J Antimicrob Chemother 2020;75:3230–8. https://doi.org/10.1093/jac/dkaa333.

[18] Mabey D, Peeling RW, Ustianowski A, Perkins MD. Diagnostics for the developing world. Nat Rev Microbiol 2004;2:231–40. https://doi.org/10.1038/nrmicro841.

[19] Soo YT, Waled SNMB, Ng S, Peh YH, Chew KL. Evaluation of EUCAST rapid antimicrobial susceptibility testing (RAST) directly from blood culture bottles. European Journal of Clinical Microbiology and Infectious Diseases 2020;39:993–8. https://doi.org/10.1007/s10096-020-03815-w.

[20] Berinson B, Olearo F, Both A, Brossmann N, Christner M, Aepfelbacher M, et al. EUCAST rapid antimicrobial susceptibility testing (RAST): analytical performance and impact on patient management. J Antimicrob Chemother 2021;76:1332–8. https://doi.org/10.1093/jac/dkab026.

[21] Jasuja JK, Zimmermann S, Burckhardt I. Evaluation of EUCAST rapid antimicrobial susceptibility testing (RAST) for positive blood cultures in clinical practice using a total lab automation. European Journal of Clinical Microbiology & Infectious Diseases : Official Publication of the European Society of Clinical Microbiology 2020;39:1305–13. https://doi.org/10.1007/s10096-020-03846-3.

[22] Tayşi MR, Şentürk GÇ, Çalişkan E, Öcal D, Miroglu G, Şencan İ. Implementation of the EUCAST rapid antimicrobial susceptibility test (RAST) directly from positive blood culture bottles without the advanced identification systems. J Antimicrob Chemother 2022. https://doi.org/10.1093/jac/dkac003.

[23] Lemos TC, Cogo LL, Maestri AC, Hadad M, Nogueira S. Short Communication Is it possible to perform bacterial identification and antimicrobial susceptibility testing with a positive blood culture bottle for quick diagnosis of bloodstream infections ? 2018;51:215–8. https://doi.org/10.1590/0037-8682-0311-2017.

